# Effect of evacuation of Japanese residents from Wuhan, China, on preventing transmission of novel coronavirus infection: a modelling study

**DOI:** 10.1101/2020.06.17.20133520

**Authors:** Yusuke Asai, Shinya Tsuzuki, Satoshi Kutsuna, Kayoko Hayakawa, Norio Ohmagari

## Abstract

**Objective:** In late January 2020, the Japanese government carried out three evacuations by aircraft from Wuhan, China, to avoid further cases of coronavirus disease 2019 (COVID-19) among Wuhan’s Japanese residents. Evacuation by aircraft may be an effective countermeasure against outbreaks of infectious diseases, but evidence of its effect is scarce. This study estimated how many COVID-19 cases were prevented among the Japanese residents of Wuhan by the evacuation countermeasure.

**Results:** Eleven imported COVID-19 cases were reported on Feb 1 from among the total 566 evacuees who returned to Japan. In the case of no evacuations being made, the cumulative number of COVID-19 cases among Wuhan’s Japanese residents was estimated to reach 25 (95% CI [20, 29]) on Feb 8 and 34 (95% CI [28, 40]) on Feb 15. A 1-week delay in the evacuation might be led to 14 additional cases and a 2-week delay to 23 additional cases. Evacuation by aircraft can contribute substantially to reducing the number of infected cases in the initial stage of the outbreak.

## Introduction

Starting at the end of December 2019, Wuhan city in China reported an increasing number of cases of coronavirus disease 2019 (COVID-2019) infection [1, 2]. Japan’s Ministry of Health, Labour and Welfare (MHLW) decided to deploy charter flights to evacuate Japanese residents of Wuhan over 3 consecutive days, on Jan 29, 30, and 31 [3]. This marked the first time that the Japanese government has carried out evacuations as a countermeasure against an infectious disease outbreak. Evacuation by aircraft may be an effective countermeasure to reduce the number of infected cases during an outbreak, but there is little quantitative evidence of its effect. Thus, the main objective of this study was to elucidate the effect of such intervention quantitatively.

## Main text

### Data sources

For this modelling study, we used data on the number of COVID-19 cases confirmed in Hubei Province for the period Jan 10-19, 2020 from a report by Wu et al. [4] and for the period Jan 20-Feb 16, 2020 from the World Health Organization (WHO) situation reports [2]. We excluded data after Feb 17 due to a change in case definition.

We collected pharyngeal swab specimens to determine the number of COVID-19 cases among the Japanese evacuees who returned on the chartered flights. The 536 evacuees answered a questionnaire about their symptoms and contact history and provided the pharyngeal swab specimens for PCR testing at that time.

### Model structure

We applied the following

susceptible-exposed-moderate-asymptomatic-infectious-recovered (SEMAIR) model to simulate the epidemic in Hubei Province:

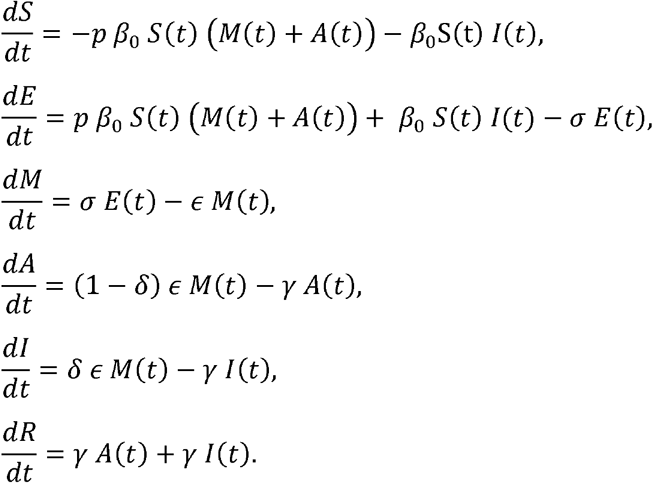

We created the model based on the assumptions that a substantial proportion of patients are asymptomatic or pauci-symptomatic (as suggested by the clinical characteristics of COVID-19) [5–7] and that transmission may be possible in mild cases (which include asymptomatic and pauci-symptomatic ones) as well as moderate cases) [5, 8, 9], but only some cases become critically ill. Hence we additionally introduced “moderate” compartment M and “asymptomatic” compartment A into the standard susceptible-exposed-infectious-recovered (SEIR) model [10, 11] to create the abovementioned SEMAIR structure. The model structure is shown in Figure 1 and detailed description of each parameter is in Appendix.

**Figure 1.**
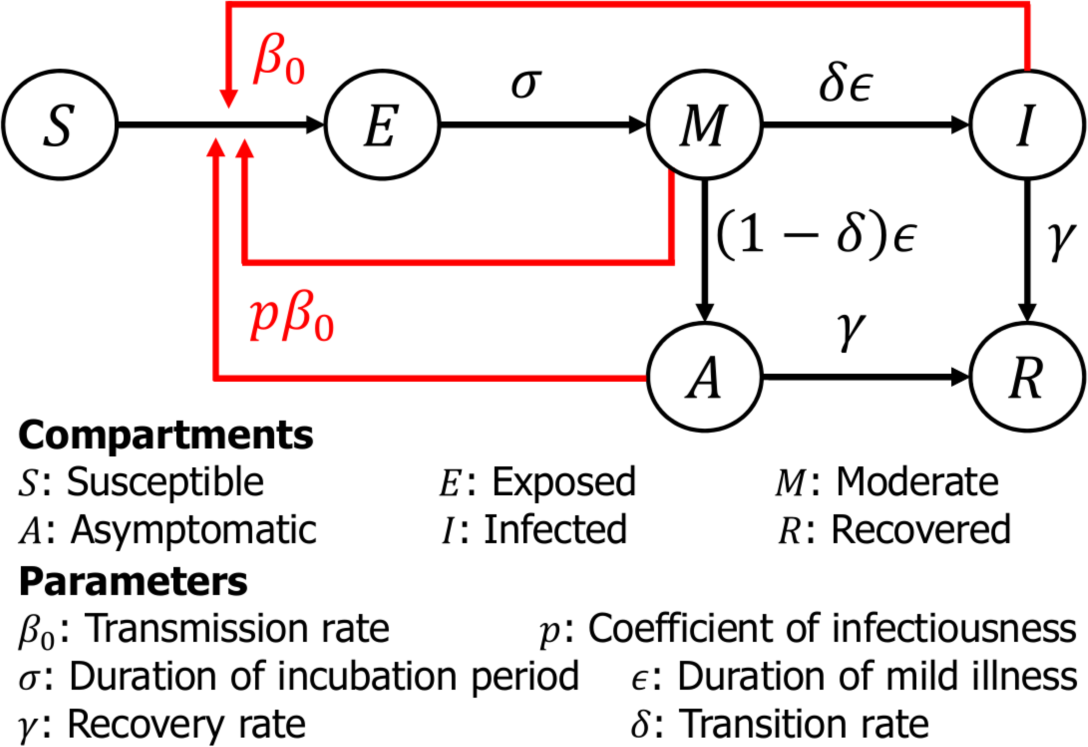
Model structure. White boxes show each compartment S (Susceptible), E (Exposed), M (Moderate), A (Asymptomatic), I (Infected), and R (Recovered) and arrows represent movement of each individual.

In the early phase of an outbreak when it is not yet recognised as such, the diagnosis rate is considered to be small and not all cases are detectable [4, 12–14]. The diagnosis rate will increase over the course of a pandemic, so we introduce time-dependent function *a*(*t*) to represent each day’s ascertainment rate, which reflects people’s awareness of COVID-19. Here we assume that *a*(*t*) is a logarithmic growth curve and that the number of reported cases *C*(*t*) is given by

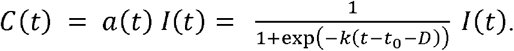

In addition to the change in diagnosis rate over time, people pay more attention to prevention and start wearing masks and washing their hands more frequently as public awareness increases. It is reported that such behavioural changes lower transmission rate *β*_0_ and this effect is captured by

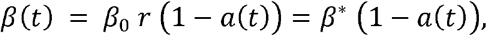

where *r* is a proportionality constant and *β*^*^ = *β*_0_ *r*.

## Results

Parameter values were randomly generated based on the estimated parameter values and their 95% CIs, and 10,000 numerical simulations were carried out to obtain the upper and lower bounds for the total number of COVID-19 patients in Hubei Province. The estimated parameter values are shown in Table 1.

**Table 1.** Values of parameters estimated in SEMAIR model for the Wuhan and Hubei population.

| | Transmission<br>rate: $\beta^*$ | Transition rate<br>from $M$ to $I$:<br>$\delta$ | Growth rate<br>of logistic<br>curve: $k$ | Time when<br>$a(t) = 1/2$:<br>$D$ | Coefficient of<br>infectiousness for<br>$M$ and $A$: $p$ |
| --- | --- | --- | --- | --- | --- |
| Wuhan | 3.11 | 0.046 | 0.34 | 13.38 | 0.73 |
| population | [3.00,<br>3.22] | [0.029,<br>0.063] | [0.32,<br>0.35] | [12.77,<br>13.99] | [0.63,<br>0.82] |
| Hubei | 3.10 | 0.047 | 0.34 | 13.40 | 0.72 |
| population | [3.09,<br>3.11] | [0.038,<br>0.056] | [0.33,<br>0.35] | [13.40,<br>14.41] | [0.68,<br>0.76] |

Estimated number of cases by SEMAIR model is shown in Figure 2. In lower left small panels, the brown dots show the total number of COVID-19 cases and the pink dots and shaded region show the 95% CI for Wuhan (612,475 [397,256, 827,695]) and Hubei Province (590,873 [487,794, 693,951]) as of Feb 16, 2020. The estimated parameters were slightly different between the Wuhan population-based analysis and the Hubei population-based analysis and the 95% CI was wider for the former than the latter, but the estimated total number of COVID-19 patients agreed well between them.

**Figure 2.**
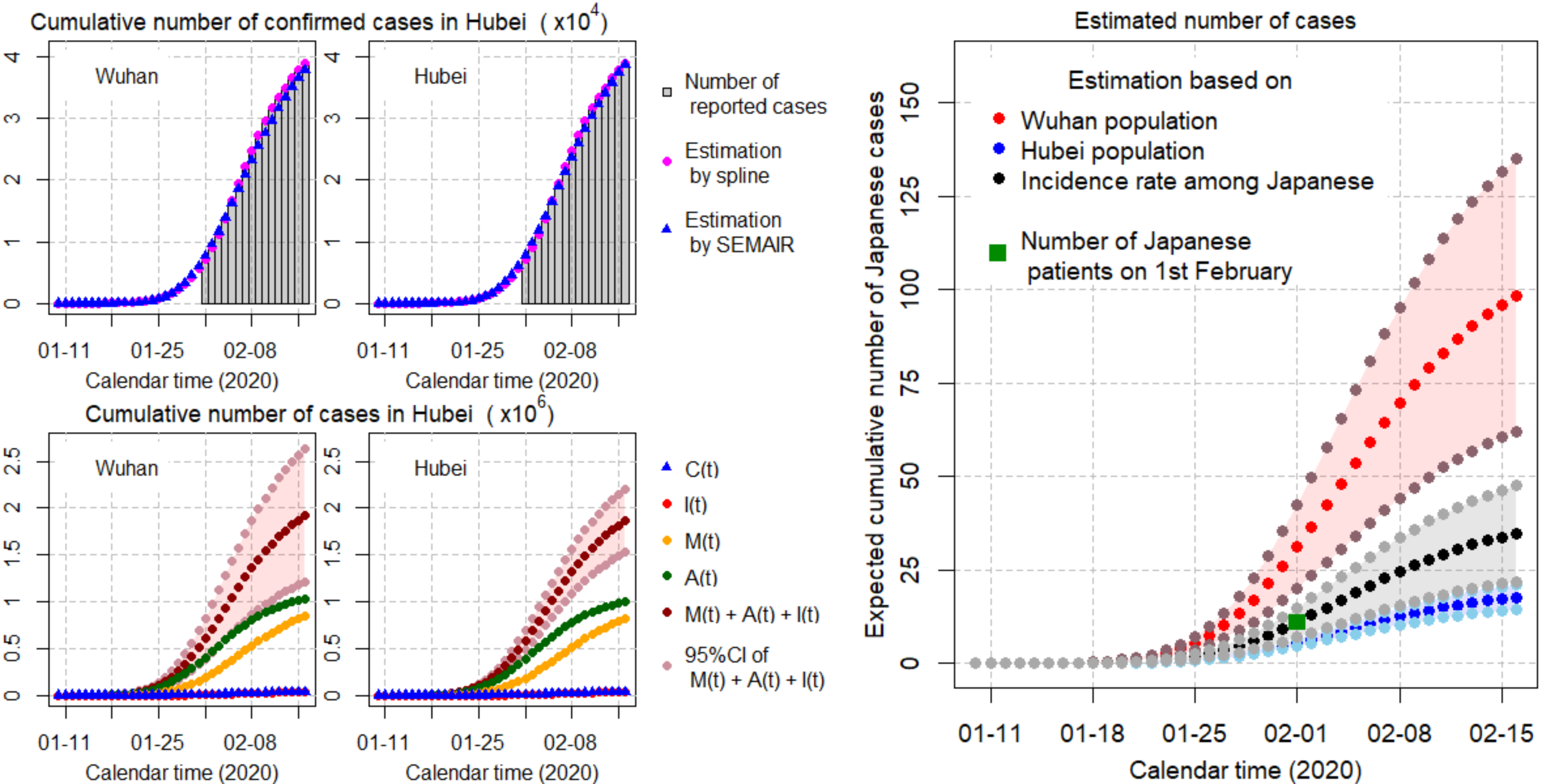
Total number of COVID-19 patients estimated by the SEMAIR model and estimated cumulative number of Japanese cases in Wuhan if the evacuations had not been carried out. The four left small panels show the number of patients estimated by the SEMAIR model. The left two panels show the estimation of cumulative number of confirmed cases and number of individuals in each compartment based on the Wuhan population. The right two panels show the estimation of cumulative number of confirmed cases and number of individuals in each compartment based on the Hubei Province population. Bar plots on the upper panels show the reported number of confirmed cases and dots and triangles show the estimation by spline method and SEMAIR model, respectively. Blue triangles and brown dots on the lower panels represent the estimated number of reported cases and the total number of cases (reported and unreported), respectively. Pink dots and the shaded region represent the 95% confidence intervals. The right large panel shows expected cumulative number of Japanese cases in Wuhan. Green square represents the observed number of Japanese patients on Feb 1. The shaded region represents the 95% confidence intervals.

Incidence rates in the Wuhan and Hubei populations were estimated as 612,475/(1.1 × 10^7^) and 590,873/(5.9 × 10^7^), respectively, and the estimated numbers of cases [95% CI] among Japanese evacuees as of Feb 1 were 31 [20, 42] and 6 [5, 7], respectively, which give bounds for over- and underestimation of Japanese evacuee cases. The actual number of cases among Japanese evacuees was 11, which lies between these estimates. The incidence rate among Japanese evacuees was 0.0195 = 11/564. By applying this rate to the Wuhan and Hubei population-based estimations, cumulative numbers of COVID-19 cases among Wuhan’s Japanese residents of were estimated as 25 [16, 33] and 25 [20, 29] on Feb 8 and as 34 [22, 46] and 34 [28, 40] on Feb 15, respectively, if the evacuation had not been carried out. As of Feb 1, there were 11 diagnosed cases among the evacuees, so a 1-week delay in the evacuation is estimated to lead to 14 additional cases and a 2-week delay to 23 additional cases. The details of estimated number of cases among Wuhan’s Japanese residents are shown in the right panel of Figure 2.

### Sensitivity analysis

We conducted sensitivity analyses for fixed parameters to reflect uncertainty. The mean values always stayed within 30 to 40 cases and details are shown in Appendix.

## Discussion

To our knowledge, this is the first study to investigate the effect of evacuation by aircraft as a countermeasure against an emerging infectious disease outbreak. Our results showed that evacuation by aircraft can contribute substantially in the short term to reducing the total cumulative number of patients.

One of the strengths of this study is that we could use all test results from the evacuees, including actual prevalence. It is noteworthy that six of 11 PCR-positive cases were asymptomatic. Furthermore, most of these 11 cases had no obvious contact history with patients with COVID-19 or influenza-like illness. These characteristics are compatible with the long duration of comparatively mild illnesses and a large number of unreported cases [5, 7, 8, 14–19]. Given this information, we modified the existing SEIR model to a SEMAIR model in order to reflect the characteristics of COVID-19.

In conclusion, evacuation by aircraft might be an effective countermeasure against an emerging infectious disease outbreak. However, further investigation is needed to assess the impact of this countermeasure more precisely.

## Limitations

We recognise several limitations of our analysis. First, we assumed homogeneity between Wuhan’s Japanese and Chinese residents, but presumably the two populations have some differences in lifestyle and behaviour and if that is the case, frequency of effective contact would be different. A previous study showed that Japanese have more frequent social contact than people in European countries [20, 21], but there is no evidence on social contact in China in the literature. Accordingly, both overestimation and underestimation should be considered when interpreting our results. Second, we fixed some of the parameters in accordance with a previous modelling study. Because these parameter values vary due to each model’s assumption, it is not clear whether fixed parameter values appropriately reflect reality or not. Nevertheless, the results of the sensitivity analyses indicate the validity of our analyses. Third, it is difficult to assess true “effectiveness” of evacuation as a countermeasure against a COVID-19 outbreak. While the number of cases averted by the intervention can be estimated, the impact of the intervention on public health is still not apparent. In addition, the Japanese government restricted the movement of evacuees after their check-up at National Center for Global Health and Medicine (NCGM), and this is thought to have prevented the spread of COVID-19 inside Japan. Restriction of movement is another effect of evacuation, but our estimation did not take it into consideration. This might be another topic appropriate for modelling studies in the future.

## Data Availability

The datasets used and/or analysed during the current study are available from the corresponding author on reasonable request.

## Abbreviations

COVID-19: coronavirus disease 2019
MHLW: Ministry of Health, Labour and Welfare Japan
WHO: World Health Organization
NCGM: National Center for Global Health and Medicine, Tokyo, Japan
SEMAIR: susceptible-exposed-moderate-asymptomatic-infectious-recovered
SEIR: susceptible-exposed-infectious-recovered
CI: confidence interval.

## Declarations

### Ethics approval and consent to participate

This study was approved by the Ethics Committee of National Center for Global Health and Medicine, Tokyo, Japan (Approval Number NCGM-G-003475-00).

### Consent for publication

Not applicable.

### Authors’ contributions

ST and YA conceived the study. YA and ST constructed the model and ran simulations. ST drafted the first manuscript and YA modified it. SK, KH and ST collected a part of data used in this study. SK, KH, and NO critically reviewed the manuscript and all authors approved the final version of the manuscript.

## Acknowledgments

Not applicable.

